# Longitudinal Analysis of Humoral and Cellular Immune Response Following SARS-CoV-2 Vaccination Supports Utilizing Point-Of-Care Tests to Enhance COVID-19 Booster Uptake

**DOI:** 10.1101/2023.04.03.23287498

**Authors:** Michael Mallory, Jennifer E. Munt, Tara M. Narowski, Izabella Castillo, Edwing Cuadra, Nora Pisanic, Paul Fields, John M. Powers, Alexandria Dickson, Rohan Harris, Richard Wargowsky, Seamus Moran, Ahmed Allabban, Kristin Raphel, Timothy A. McCaffrey, James D. Brien, Christopher D. Heaney, John E. Lafleur, Ralph S. Baric, Lakshmanane Premkumar

**Affiliations:** Department of Microbiology & Immunology, University of North Carolina School of Medicine, Chapel Hill, North Carolina, USA; Department of Epidemiology, UNC Chapel Hill School of Public Health, University of North Carolina at Chapel Hill, Chapel Hill, North Carolina, USA; Department of Environmental Health and Engineering, Johns Hopkins Bloomberg School of Public Health, Baltimore, Maryland, USA; Adaptive Biotechnologies, Seattle, Washington, USA; Department of Molecular Microbiology & Immunology, Saint Louis University School of Medicine, Saint Louis, Missouri, USA; Department Emergency Medicine, George Washington University School of Medicine, Washington, DC, USA; Department of International Health, Bloomberg School of Public Health, Johns Hopkins University, Baltimore, Maryland, USA

## Abstract

Individuals with weaker neutralizing responses show reduced protection with SARS-CoV-2 variants. Booster vaccines are recommended for vaccinated individuals, but the uptake is low. We present the feasibility of utilizing point-of-care tests (POCT) to support evidence-based decision-making around COVID-19 booster vaccinations. Using infectious virus neutralization, ACE2 blocking, spike binding, and TCR sequencing assays, we investigated the dynamics of changes in the breadth and depth of blood and salivary antibodies as well as T-cell clonal response following mRNA vaccination in a cohort of healthcare providers. We evaluated the accuracy of two POCTs utilizing either blood or saliva to identify those in whom humoral immunity was inadequate. >4 months after two doses of mRNA vaccine, SARS-CoV-2 binding and neutralizing Abs (nAbs) and T-cell clones declined 40-80%, and 2/3rd lacked Omicron nAbs. After the third mRNA booster, binding and neutralizing Abs increased overall in the systemic compartment; notably, individuals with previously weak nAbs gained sharply. The third dose failed to stimulate secretory IgA, but salivary IgG closely tracked systemic IgG levels. Vaccine boosting increased Ab breadth against a divergent bat sarbecovirus, SHC014, although the TCR-beta sequence breadth was unchanged. Post 3rd booster dose, Ab avidity increased for the Wuhan and Delta strains, while avidity against Omicron and SHC014 increased to levels seen for Wuhan after the second dose. Negative results on POCTs strongly correlated with a lack of functional humoral immunity. The third booster dose helps vaccinees gain depth and breadth of systemic Abs against evolving SARS-CoV-2 and related viruses. Our findings show that POCTs are useful and easy-to-access tools to inform inadequate humoral immunity accurately. POCTs designed to match the circulating variants can help individuals with booster vaccine decisions and could serve as a population-level screening platform to preserve herd immunity.

**One Sentence Summary:** SARS-CoV-2 point-of-care antibody tests are valuable and easy-to-access tools to inform inadequate humoral immunity and to support informed decision-making regarding the current and future booster vaccination.

## Introduction

The original SARS-CoV-2 vaccines developed by Moderna (mRNA-1273) and Pfizer/BioNTech (BNT162b2) utilize a synthetic messenger RNA (mRNA) coding for the Wuhan strain spike protein^1, 2^. After 2-dose mRNA vaccination, neutralizing antibody (nAb) levels varied among naïve-vaccinated (NV) and infected-vaccinated (IV) individuals^3, 4^, and were typically less effective against SARS-CoV-2 variants of concern as compared to the ancestral Wuhan strain^3^. Major SARS-CoV-2 variants, such as Delta and Omicron, have accumulated mutations within the receptor binding domain (RBD) of the spike protein. These mutations alter spike protein antigenic properties and affect vaccine-induced antibody (Ab) binding and neutralizing activities^5^. Some spike RBD mutations enhanced viral interaction with the host cell-surface receptor, ACE2^6^. Importantly, individuals with weaker neutralizing responses against the Wuhan strain showed reduced protection with SARS-CoV-2 variants^7^.

As the Omicron subvariants emerged, the information about the waning immunity following vaccination caused a major concern^8, 9^. Yet, millions of eligible individuals have not opted to receive the third dose booster targeting ancestral Wuhan spike protein^10^. Though widely available, far fewer have received the newly approved bivalent booster targeting both the original Wuhan strain and Omicron sub-variants^10^. Various factors may contribute to booster hesitancy^11, 12^, including questions about booster effectiveness against newer, emerging variants. Some individuals developed vaccine-induced side effects^13–16^, which may also influence decisions about the risks/benefits of vaccinating. Thus, strategies that grow vaccine confidence and encourage vaccine uptake are urgently required to protect individuals with weakened immunity against evolving SARS-CoV-2 strains.

Our previous study of mRNA vaccine-induced Ab responses in a healthcare providers (HCP) cohort showed the development of variable neutralizing antibodies in IV and NV subjects^3^. In this current manuscript, we explored a strategy to identify individuals with weak vaccine responses. We investigated the breadth and depth of blood and salivary Abs and T-cell clonal response before and after the original third booster dose. We also tested the ability of two lateral-flow, low-cost point-of-care tests (POCTs) to inform weak functional Ab levels in both blood and salivary samples. Our results demonstrate that POCTs strongly agree with functional neutralizing Ab levels and can help evidence-informed decisions regarding the current and future booster vaccination.

## Results

### SARS-CoV-2 Ab and TCR-β clonal responses decline >4 months after the second-dose of the mRNA vaccine

To evaluate the levels of binding and functional Abs after two-dose mRNA vaccination, we analyzed samples collected at two-time points post-second dose: T2a (4-9 weeks) and T2b (19-36 weeks) from 35 naïve-vaccinated (NV) and 9 infected-vaccinated (IV) subjects in the HCP cohort. We measured spike and RBD binding, RBD-ACE2 blocking Ab activity, and infectious virus neutralization towards the Wuhan strain using the methods previously established^3, 17^ (Figure 1). Spike and RBD binding Ab levels declined 43-77% between T2a and T2b in both the NV and IV subjects (Figure 1, A-D). RBD-directed Abs decreased more than overall spike-directed Abs, regardless of NV or IV status. NV subjects also showed a comparable reduction in ACE2 blocking Abs (Figure 1, E) and SARS-CoV-2 neutralizing activities, 62% and 56%, respectively (Figure 1, F). However, the decline among IV subjects was more modest for ACE2 blocking Ab (33% change, p=0.0332) (Figure 1, G) and insignificant for nAbs (7% change, Figure 1, H) during this period.

**Figure 1:**
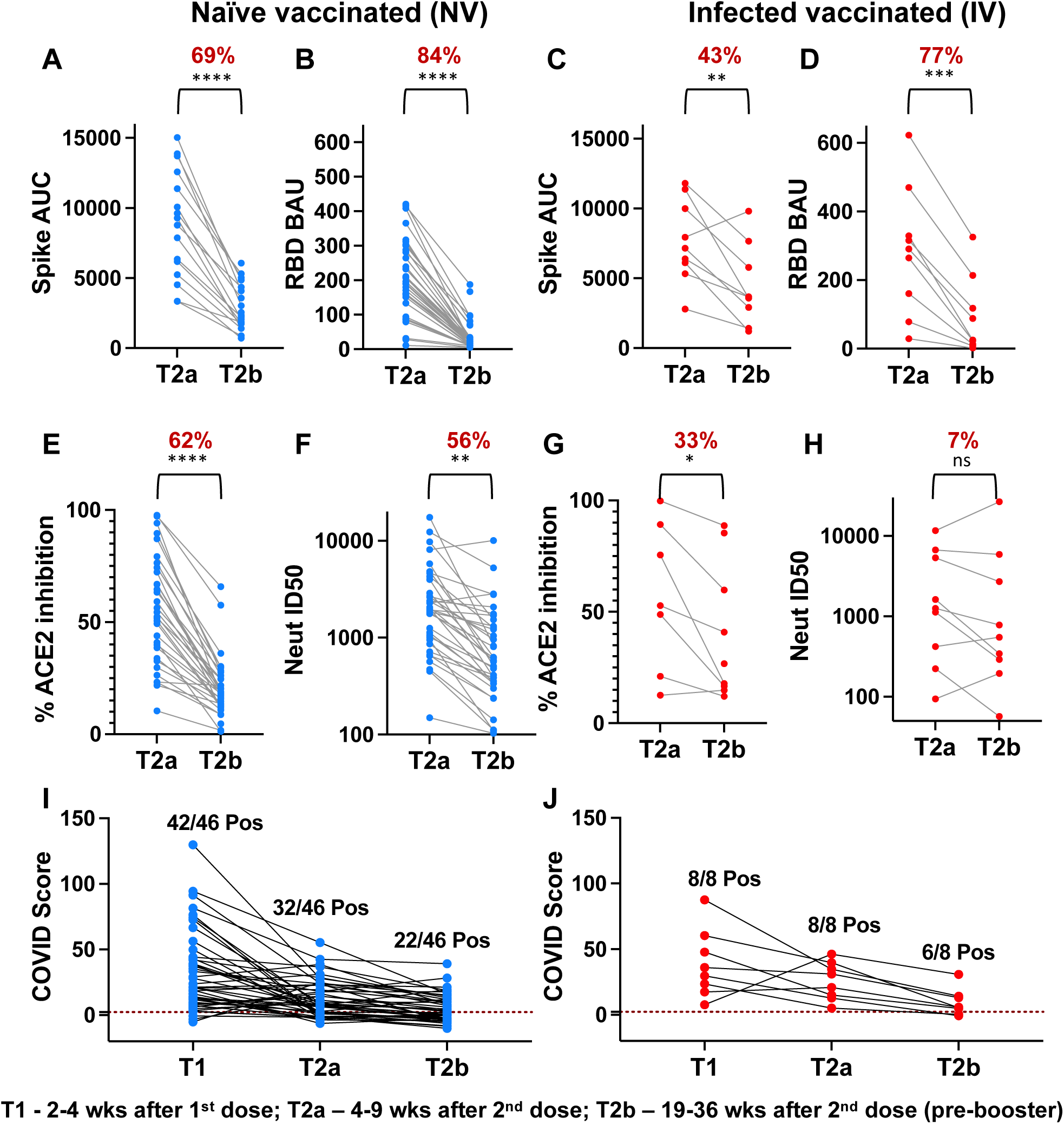
Dynamics of Ab and T-cell clonal responses to Wuhan strain after mRNA vaccine. Analysis of Spike (A and C) or RBD (B and D) binding Abs, ACE2 blocking Abs (E and G), and neutralizing Abs (F and H) to the Wuhan strain between 4-9 weeks (T2a) and 19-36 weeks (T2b) after dose-2. Naïve vaccinated (NV) subjects (n=35) and infected vaccinated (IV) subjects (n=9) are shown in blue and red dots, respectively. The mean percent decrease in Ab levels and paired t-test summary (0.12 (ns), 0.0332 (*), 0.0021 (**), 0.0002 (***), <0.0001 (****)) between the two time points are shown in brackets above the plot. Longitudinal analysis of T-cell clonal response at 2-4 weeks after dose-1 (T1), 4-9 weeks after dose-2 (T2a), and 19-36 weeks after dose-2 (T2b) in (I) NV (n=46, blue) and (J) IV subjects (n=8, red). A COVID score threshold of >2.23 was applied to classify the subjects as positive or negative. Percent positivity at each time-point is shown.

Next, to measure the dynamics of the cellular response, we conducted immunosequencing of the CDR3 regions of human TCRβ chains for 46 NV and 8 IV subjects. We applied a statistical classifier (COVID-Score), previously established for SARS-CoV-2^18, 19^, that demonstrated high specificity and sensitivity with the samples collected at baseline (T0) and after the first dose of the mRNA vaccine (T1) within the HCP Cohort (Supplementary Figure 1). The COVID score declined between T1 and T2a sampling points, and by T2b, the COVID-Score fell below the assay positive threshold (COVID-Score=2.23) for 24 of 46 NV and 2 of 8 IV subjects (Figure 1, I and J). These data suggest that the mRNA vaccine-induced Ab and cellular clonal responses decline significantly >4 months following the second dose in our HCPs cohort. However, the declining trend observed between T1 and T2a for the cellular clonal response is quite the opposite of the direction we reported previously for humoral response in this cohort^3^.

### Circulating SARS-CoV-2 Abs and TCR-β clones increase after the third booster dose of the mRNA vaccine

To compare the levels of Ab and cellular responses before and after the third booster dose (Wuhan), we collected additional samples from 33 subjects in the HCP cohort around 3-18 weeks post-third dose (T3). We measured RBD Ab binding and live-virus Ab neutralization directed at the Wuhan, Delta (B.1.617.2), and Omicron (B.1.1.529/BA.1) strains and compared their levels between T2b and T3 sampling time points (Figure 2A and 2B). At T3, depending on the variant tested, the mean RBD binding activity increased 6-9 fold, whereas neutralizing activities rose 17-49 fold (p<0.0001). Notably, RBD binding against the Omicron variant was low, with 21 of 33 subjects lacking detectable levels of Omicron neutralizing activity before the third dose. We then measured ACE2 blocking activities against a panel of SARS-CoV-2 strains, including Omicron sub-variants, to assess the contribution of RBD-targeting Abs in the humoral repertoire following SARS-CoV-2 in IV and NV combined (Figure 2C). The increase in ACE2 blocking activity following booster dose was consistent with the increase in RBD binding activities across several variants (Fig 2A and Fig 2C). The smallest change in %ACE2 blocking activity was towards Omicron subvariant BA.3, followed by the Wuhan and Alpha strains, though T2b %ACE2 blockage was highest across the board towards Wuhan and Alpha, perhaps contributing to the minor fold change seen between timepoints. However, %ACE2 blockage in the Omicron subvariants was more dependent on the individual, with differences between T2b to T3 being less significant (p=ns-0.0021) than for all other variants (p<0.001). In contrast to the declining trend observed between the first and second doses, the third booster dose stimulated the T-cell clonal response about 3-fold (p<0.0001, Figure 2D).

**Figure 2:**
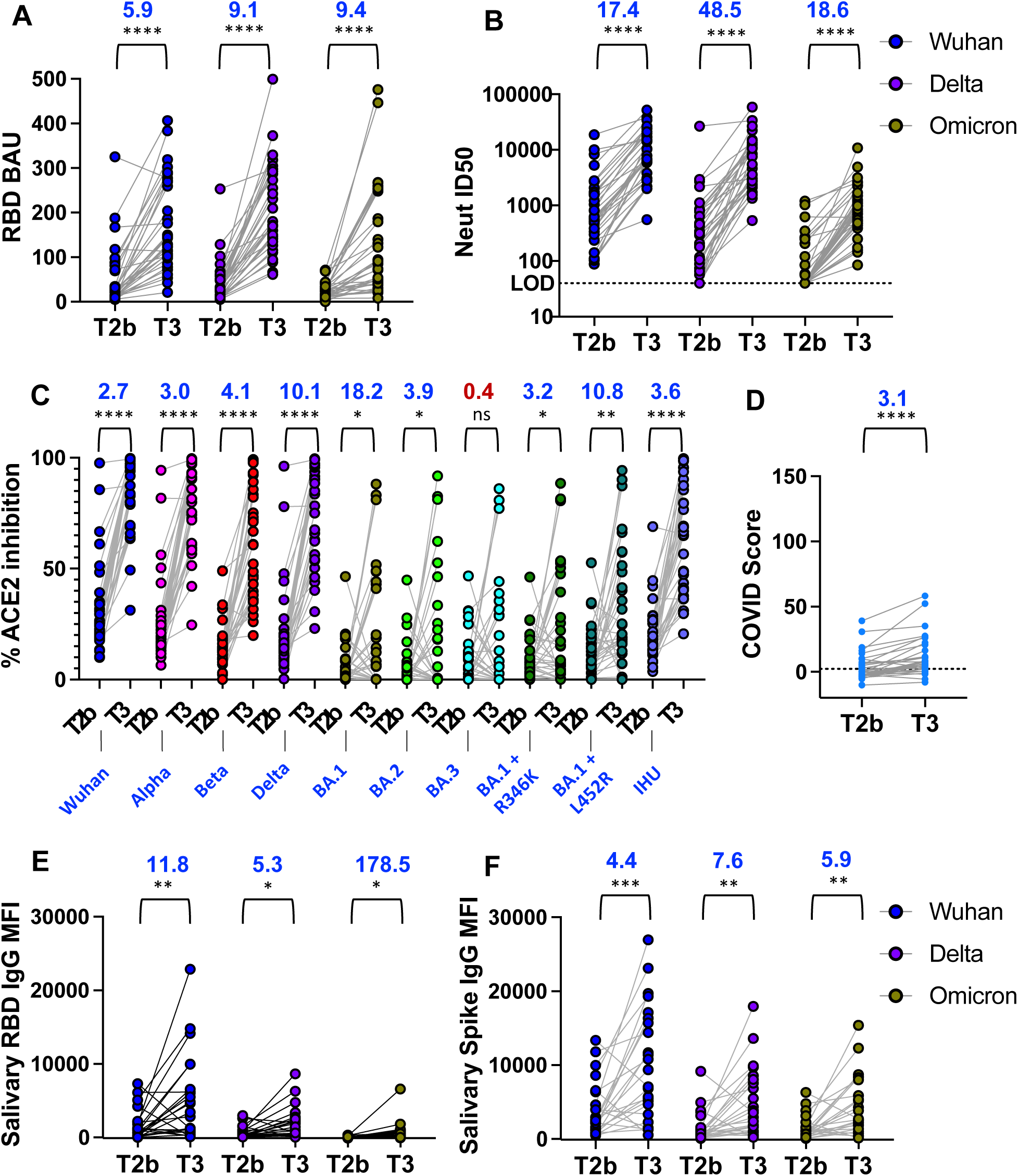
Dynamics of Ab and T-cell clonal responses after the third dose of the mRNA vaccine. Analysis of (A) RBD binding Abs, (B) neutralizing Abs, and (C) ACE2 blocking Abs to the Wuhan and other variants in blood samples in 33 subjects between 19-36 weeks after dose-2 (T2b) and 3-18 weeks after dose-3 (T3). (D) Analysis of SARS-CoV-2 TCR-β levels before and after dose-3 in blood samples from 33 subjects. The threshold for TCR sequencing assay (2.23) is shown as a dotted line. Analysis of (E) RBD and (F) Spike binding Abs to the Wuhan and other variants in salivary samples from 24 subjects at T2b and T3 time points. The mean fold change in SARS-CoV-2 Abs or TCR-β genes and paired t-test summary (0.12 (ns), 0.0332 (*), 0.0021 (**), 0.0002 (***), <0.0001 (****)) between the two-time points in blood and salivary samples are shown in brackets above the plot. Wuhan (blue), delta (violet), and Omicron BA1 (bottle green). Other SARS-CoV-2 variants tested in panel C are labeled below the plot.

### Vaccine boosters and mucosal immunity in saliva

To study whether mRNA vaccines induce mucosal immunity following the third booster dose, we assessed the development of Ab responses to spike and RBD with the saliva samples collected at T2b and T3 time points (Figure 2, E, and F). RBD IgG levels were low for Wuhan and Delta strains, and 3/4th of the participants had undetectable levels of RBD IgG for Omicron strains before the booster (T2b). Spike-directed IgG binding was higher at T2b compared to RBD-directed IgG; however, both Ab types increased significantly at T3 (p=0.0332-0.0002), though we observed no evidence for induction of salivary secretory IgA (sIgA) after the third booster (Supplementary Figure 2). Together these data suggest systemic and salivary spike IgG Abs and T-cell clonal response increase following the third booster dose.

### Booster-mediated increase in SARS-Co-2 Ab and T-cell clonal response applies across heterogeneous groups of subjects

We previously reported heterogenous nAb responses following the first and second doses of mRNA vaccines in our HCP cohort. We observed that nAb response fell into three distinct patterns: Group-I (G-I) had a weak response overall; Group-II (G-II) had a strong reaction with subsequent decline; Group III (G-III) had a strong response which remained strong. In this current study, we tested RBD binding and live-virus neutralization in these three groups across the Wuhan, Delta and Omicron variants at three timepoints: T2a, T2b, and T3. Across all groups and variants, RBD binding and nAbs declined between T2a and T2b time points (Figure 3, A-F, Supplementary Figure 3). The third booster dose (T3) stimulated RBD binding and nAbs for all strains. However, the mean fold increase in RBD binding and neutralizing Abs levels were higher for G-I than for G-II and G-III. Before the booster, G-I had the lowest median RBD binding and neutralizing Abs to the Wuhan and Delta strains while its Omicron neutralization titers were below the assay detection limit (Supplementary Fig 3). By comparison, G-I binding and neutralizing Ab levels were 2-3 fold lower than G-II; Likewise, G-II was 2-3 fold lower than G-III. After the third booster (T3), G-I and G-II RBD binding and neutralizing Abs were comparable, while G-III remained higher (Supplementary Figure 3).

**Figure 3:**
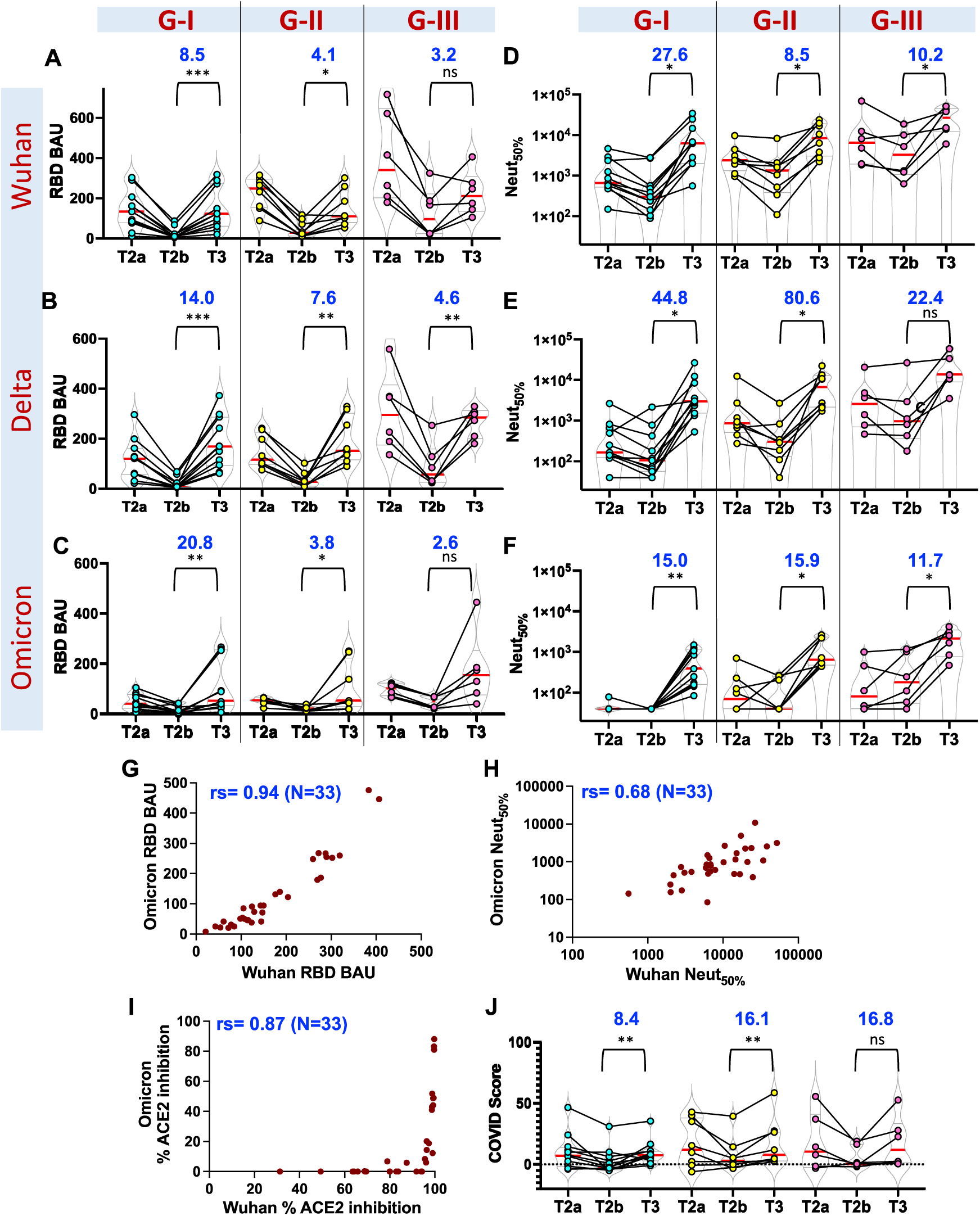
Dynamics of Ab and T-cell clonal responses before and after the third dose in heterogeneous groups. Analysis of (A-C) RBD binding and (D-F) neutralizing Abs to the Wuhan, Delta, and Omicron variants in different subject groups at 4-9 weeks after dose-2 (T2a), 19-36 weeks after dose-2 (T2b) and 3-18 weeks after dose-3 (T3). Three groups were previously classified based on the patterns of the neutralizing Ab response. A subset is analyzed in this study: Group I had a weak neutralizing Ab response after complete vaccination. Group II had a robust neutralizing Ab response after dose-1 that declined after dose-2. Group III neutralizing Abs rose after dose-1 and after dose-2. Correlation plots for (G) RBD binding, (H) neutralizing, and (I) ACE-2 blocking Ab levels between Wuhan and Omicron strain. The non-parametric Spearman correlation coefficient (rs) for each plot is shown. (J) Analysis of SARS-CoV-2 TCR-β levels at T2a, T2b and T3 sampling points. The threshold for TCR sequencing assay (2.23) is shown as a dotted line. The mean fold change in SARS-CoV-2 Abs or TCR-β genes and paired t-test summary (0.12 (ns), 0.0332 (*), 0.0021 (**), 0.0002 (***), <0.0001 (****)) before and after the booster in Group I (n=11), Group II (n=8), and Group III (n=6) are shown in brackets above the plot.

Remarkably, individuals with high levels of RBD binding, ACE2 blocking, and live-virus neutralizing activities against the Wuhan strain also had higher activities against the Omicron strain (Figure 3, G-to-I). COVID-Score also increased significantly from T2b to T3 for G-I and II groups (Figure 3J), suggesting that the booster dose improved both Ab and cellular clonal responses for most participants. Importantly, the low responders appear to benefit the most from the booster mRNA vaccine.

### After the third booster dose, the avidity and breadth of the Abs grew, but not the TCR breadth

Next, to assess the overall strength of RBD-directed antibodies developed after the booster, we measured avidity (“functional affinity”) before (T2a and T2b) and after the third booster dose (T3) against the RBD of the Wuhan, Delta, and Omicron strains. To do this, we used a modified RBD ELISA which employs 4 M urea as a chaotropic agent to dissociate low-avidity Abs from RBD while allowing only high-avidity Abs to remain associated. The ratio of the Abs bound with and without urea was used to express the avidity index (AI) (Figure 4). Between the T2a and T2b sampling time points, the RBD Ab avidity index improved against the Wuhan and Delta strains, although the overall plasma Ab levels decreased (Figure 4A-B and G). Before the third booster dose, Omicron binding was low, rendering the avidity index indeterminate (Figure 4C and G). However, after the third booster dose, plasma Ab levels towards all strains were much higher than observed at both T2a and T2b (Figure G), while the Wuhan and Delta RBD Ab avidity index further improved to levels significantly higher than observed at the earlier time points. Strikingly, the Omicron RBD Ab avidity index and the Ab levels in plasma after the third booster dose reflected the levels observed for the Wuhan and Delta strains at T2b (Figure 4A-C and G).

**Figure 4.**
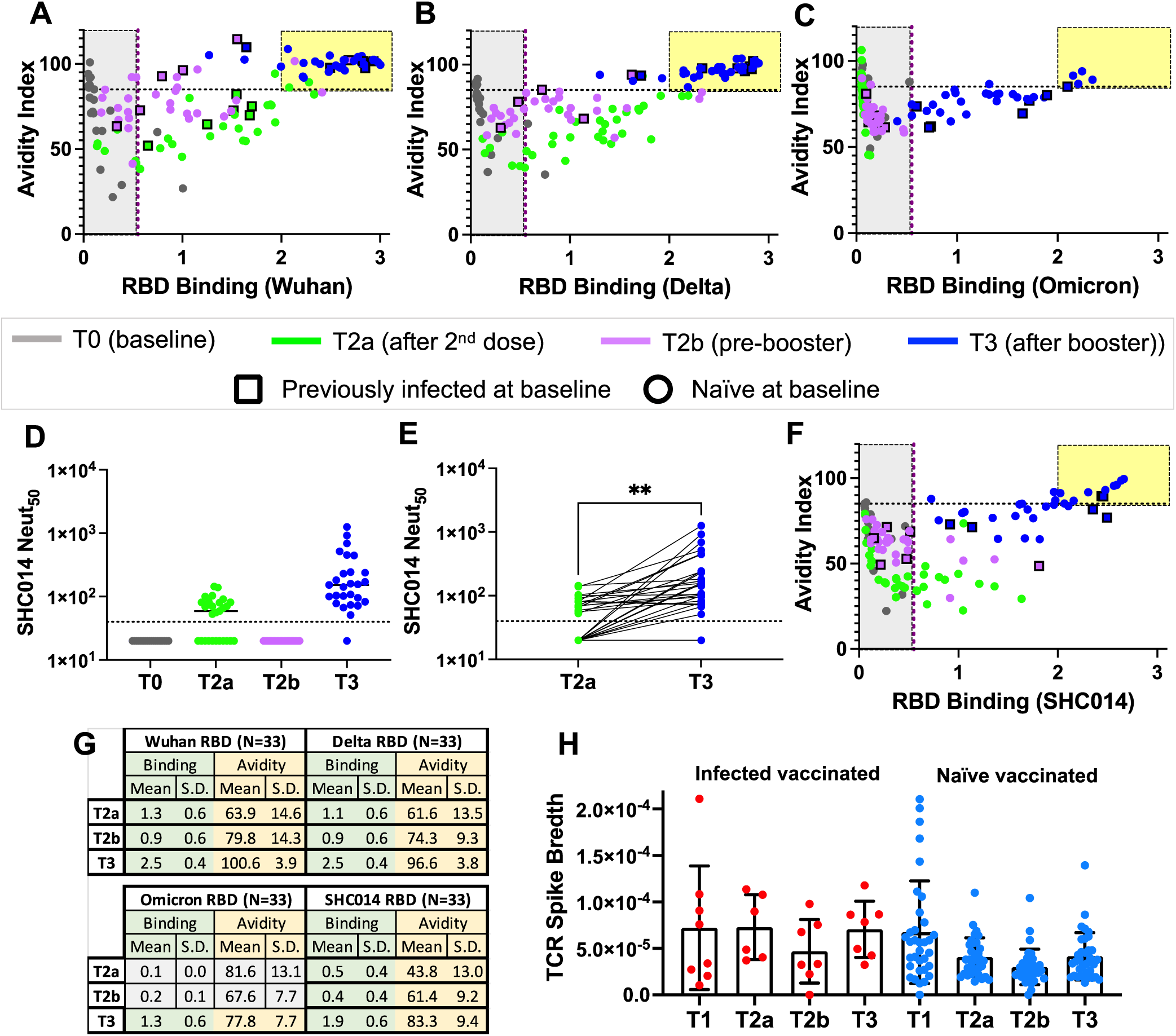
Avidity and breadth of the SARS-CoV-2 Abs and TCR-β CDR3s. Assessment of Ab binding strength following mRNA vaccine against RBD from (A) Wuhan, (B) Delta, (C) Omicron, and (F) sarbecovirus bat SHC014 strain at different time points. The avidity index, calculated by measuring Ab binding after treatment with urea solution relative to without urea treatment, has been plotted against binding without urea treatment in 33 subjects (square, previously infected at pre-vaccine; circle, naive at pre-vaccine). The yellow box indicates elevated levels of high-avidity Abs in plasma. Samples analyzed at respective pre-vaccine and post-vaccine time points (T1, 2-4 weeks after dose-1, T2a, 4-9 weeks after dose-2 (green), T2b, 19-36 weeks after dose-2 (pink), and T3, 3-18 weeks after dose-3 (blue) are shown. (D-E) Analysis of neutralizing Ab levels against sarbecovirus bat SHC014 strain at different time points as above in 27 subjects. (G) RBD Binding and avidity index summary for the four strains at three sampling time points (T2a, T2b, and T3). The mean and standard deviations for 33 subjects are summarized. For the Omicron RBD, binding is low at T2a and T2b, rendering the avidity index indeterminate (gray). (H) Evaluation of sequence diversity of SARS-CoV-2 spike TCR-**β** at different time points in 42 subjects. Naive vaccinated (n=34) and infected vaccinated (n=8) subjects are shown in blue and red dots at different time points, as indicated above. Paired t-test, 0.0021 (**).

To gain insights into the breadth of mRNA vaccine-induced Abs before (T2a and T2b) and after the third booster dose (T3), we measured neutralizing Abs to a bat SARS-like coronavirus RsSHC014 (SHC014), a phylogenetically distant Sarbecoviruses that uses the ACE2 receptor. We observed a low SHC014 neutralizing Ab titer after T2a that declined to an undetectable level at T2b (Figure 4D). After the booster, the SHC014 neutralizing Ab titer was significantly higher than at T2a (p=0.0016, Figure 4E). Next, we evaluated the RBD Ab avidity index and binding for SHC014 at T2a, T2b, and T3 time points. In agreement with the SHC014 neutralizing Ab titers, the low avidity RBD Abs developed after the second dose of the mRNA vaccine wanned by T2b (Figure 4F and G). After the booster, the RBD Ab avidity index and the Ab levels in plasma for the SHC014 were marginally better than the Wuhan RBD Abs levels at T2b and the Omicron RBD Ab levels in plasma at T3 (Figure 4G).

We have also observed the elevation of spike Abs to seasonal human coronaviruses (HCoVs) after the first dose of the SARS-CoV-2 mRNA vaccine in our cohort (Supplementary Figures 4A and 4B). However, these cross-reactive binding Abs do not contribute to the cross-neutralization, as evidenced by the neutralization titers between baseline (T0) and after the first dose (T1) for NL63 (seasonal α-HCoV) and OC43 (seasonal β-HCoV) (Supplementary Figure 4C and 4D). While we observed an enhancement of avidity and breadth in the humoral response following the third booster dose, the breadth and diversity in the SARS-CoV-2 spike-specific CDR3 regions of human TCRβ chains were unchanged in samples collected at the T3 sampling point, as compared to T2a (Figure 4H).

### Low-cost Rapid LFA blood test informs low circulating functional Abs

Even though our data suggest that boosting with the SARS-CoV-2 mRNA vaccine third dose enhances avidity and breadth of neutralizing Abs across heterogeneous subjects, only <20% of the population have opted for booster vaccination among those who have received their second dose^10^. To help build tools to support evidence-informed decision-making about the COVID-19 booster vaccination, we conducted experiments to explore simple laboratory binding assays that are widely available and can inform the lack of functional Abs in blood samples. We initially evaluated if our RBD ELISA binding results stratified by low (≤ 22 BAU) and high (>22 BAU) could predict the weak infectious live-virus neutralizing activity (Figure 5A) or ACE2 blocking activity (Figure 5B) in blood samples collected at T2b (19-36 weeks after dose 2). Remarkably, the lowest RBD binders aligned well with weak neutralizing and ACE2 blocking activities, indicated by the high statistical significance (p<0.0001).

**Figure 5.**
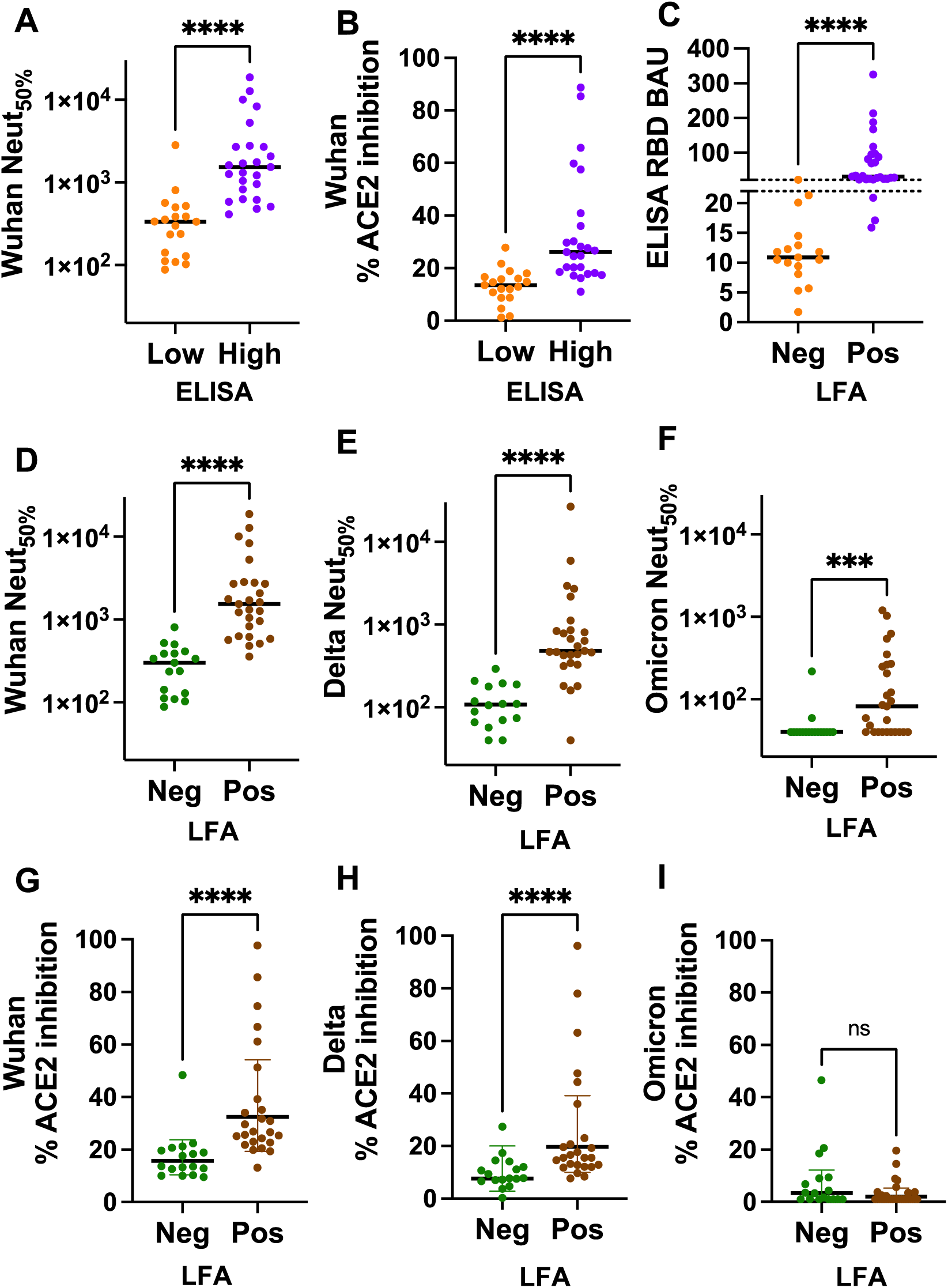
Low-cost Rapid LFA blood test to detect low circulating Abs. Assess low ELISA RBD binding agreements with the following: (A) weak neutralization, (B) weak ACE2 blocking activity, and (C) negative LFA blood test. Assess negative LFA blood test agreements with neutralizing titers of (D) Wuhan, (E) Delta, and (F) Omicron strains. Assess negative LFA blood test agreements with ACE2 blocking activities of (G) Wuhan, (H) Delta, and (I) Omicron strains. The p-value summary from the Mann-Whitney non-parametric unpaired test to determine the difference between the groups is shown above the scatterplot. ****p < 0.0001; ***p = 0.0008; ns, >0.1234.

Next, we tested if a low-cost lateral flow-based test could predict the weak live-virus neutralization and ACE2-blocking activities in blood samples. As a proof-of-concept, we evaluated the emergency use authorized Cellex™ qSARS-CoV-2 IgG/IgM Rapid test, which utilizes the RBD of the Wuhan strain and can be performed with a drop of a blood sample (∼10 µl) and read in less than 20 minutes (Supplementary Figure 5). As shown in Figure 5C, the negative test result of the Cellex Rapid test highly correlated with low RBD binding activity (p<0.0001), warranting further evaluation with functional assays. Next, we assessed if the Rapid test results, separated into negatives and positives, could inform weak neutralizing activities against the Wuhan, Delta, and Omicron strains using blood samples collected at T2b from 44 subjects. For both Wuhan and Delta strains, the negative Rapid test results were highly correlated with the weak live-virus neutralization and ACE2 blocking activities with high statistical significance (p<0.0001). The correlation between the negative Rapid test results and the Omicron live-virus neutralizing activity was also significant, although lower than the Wuhan and Delta strains (p=0.008). The Rapid test results poorly correlated with ACE2 blocking activities for Omicron, as most individuals tested had poor ACE2 blocking activities at the T2b time point.

### Low-cost Rapid LFA saliva informs low circulating functional Abs

As salivary samples are an attractive alternative to blood testing due to their non-invasive nature and ready repeatability, we assessed the correlation between the Wuhan spike and RBD binding Abs in blood and salivary samples (Figures 6A and B). Both spike and RBD IgGs from saliva and blood were strongly correlated despite being measured by two independent methods (Luminex bead assay vs. ELISA), as saliva IgG primarily derives from plasma by transudation from the gingival blood circulation^20–22^. As a proof-of-concept, we assessed the emergency use authorized CoVAb SARS-CoV-2 IgG test, which measures RBD IgG in salvia in 15 minutes (Supplementary Figure 6). The correlation between the negative test results by the CoVAb IgG test and the low RBD binding activity measured with blood aligned with high significance (p<0.0001) (Fig 6C).

**Figure 6.**
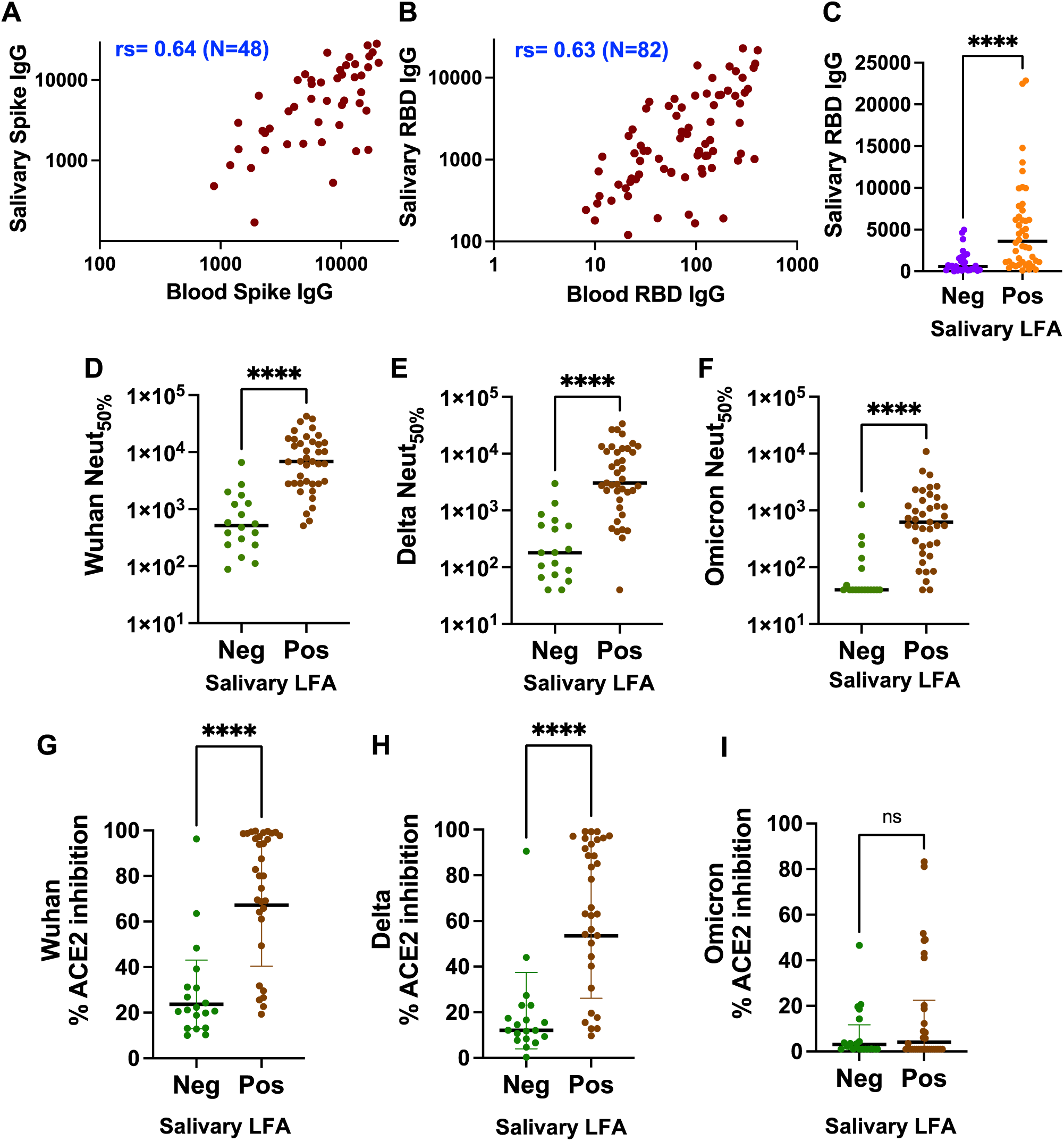
Low-cost Rapid LFA saliva test to detect low circulating Abs. Assess the correlation between blood and salivary IgG against (A) Spike and (B) RBD. The non-parametric Spearman correlation coefficients (rs) for the plots are shown. (C) Assess low Luminex RBD binding agreements with the negative LFA saliva test. Assess negative LFA salvia test agreements with neutralizing titers of (D) Wuhan, (E) Delta, and (F) Omicron strains. Assess negative LFA saliva test agreements with ACE2 blocking activities of (G) Wuhan, (H) Delta, and (I) Omicron strains. The p-value summary from the Mann-Whitney non-parametric unpaired test to determine the difference between the groups is shown above the scatterplot. ****p< 0.0001; ns, >0.1234.

The negative CovAb test results were highly correlated with the weak live-virus neutralizing and ACE2 blocking activities for the Wuhan and Delta strains with high significance (p<0.0001) (Fig 6 D, E, G, H). While the negative CovAb test results were also highly linked with the live-virus neutralizing activity for the Omicron strain (p<0.0001) (Fig 6F), the salivary LFA could not differentiate between low and high ACE2 blocking activity, which was negative for most samples tested (Fig 6 I). These results support that the lateral-flow assay platforms using saliva or blood samples can inform individuals of weak SARS-CoV-2 functional, especially neutralizing, Ab responses.

## Discussion

Antibody-mediated SARS-CoV-2 neutralization mainly occurs through recognizing conformational epitopes on the spike protein^23^. In contrast, the TCR reactivity occurs through presentation of several viral peptides displayed in the context of human leukocyte antigens on the surface of infected cells; consequently, T-cell responses are less sensitive to mutations in the spike protein^24, 25^. Human challenge studies are required to thoroughly understand the significance of humoral and cellular immunity for protection against SARS-CoV-2 infection. Nevertheless, in macaque challenge studies, neutralizing antibodies protected against SARS-CoV-2 infection in a dose-dependent manner^26^. At the same time, the cellular immune response reduced symptomatic and severe disease against SARS-CoV-2 infection in the setting of waning and sub-optimal neutralizing antibody titers^26^. A predictive model based on studies of seven different vaccines and convalescent cohorts estimated that neutralization level strongly correlates with immune protection against SARS-CoV-2 in humans^7^. Similarly, hospitalized COVID-19 patients who received high tittered (>1:640) neutralizing convalescent plasma were discharged earlier than patients who received convalescent plasma with lower nAb titers^27^. These studies support that higher systemic neutralizing antibody levels predict SARS-CoV-2 protection.

The emergence of antigenically distinct variants, such as Omicron and its subvariants that carry multiple mutations in the spike protein, has significantly reduced the effectiveness of vaccine-induced nAbs^28^. Individuals with high neutralizing antibody titers were more protected against variants than those with weak neutralizing Ab levels^3^. Yet, the protection against hospitalization and death among individuals experiencing a breakthrough infection with a SARS-CoV-2 variant was low, suggesting that T cells play a significant role against severe disease in the context of infection by the SARS-CoV-2 variant^29^. In our cohort of healthy adults, T-cell clonal levels fluctuated, while breadth of the TCRβ was unchanged after the second dose (T2a) and third dose (T3). A comparative study of a healthy adult cohort that analyzed the dynamics of T-cell stimulation to produce cytokines showed that T-cell immunity is established and maintained after complete vaccination, as the T-cell effector function remained stable before and after the booster vaccination^30^. These observations support that the booster vaccination is more crucial for improving the neutralizing antibody levels than the T-cell response^30^.

In our HCP cohort, we observed a significant decrease in nAb levels >4 months after the second dose of the mRNA vaccine, though more pronounced in the NV subjects than in the IV subjects. Individuals who developed strong nAbs against the Wuhan strain shortly after the second dose had modest nAbs against the Omicron strain six months out (T2b). However, individuals who developed inadequate nAb response after the second dose entirely lacked nAbs against the Omicron strain after ∼6 months (Supplementary Figure 3, Group I and II), suggesting that waning of immunity following vaccination is not limited to at-risk groups, including older/immunocompromised recipients^31^, but is applicable population-wide^7, 32^.

After boosting with a Wuhan strain-based mRNA vaccine, binding and neutralizing Abs sharply rose against Wuhan, Delta (B.1.617.2), and Omicron (B.1.1.529) strains for most individuals in our cohort. However, the relative gain for the subjects who had inadequate neutralizing Abs after the second dose was striking, highlighting the importance of receiving a booster dose, especially for those that did not experience a robust neutralizing response after complete two-dose vaccination. Though we did not find evidence for salivary secretory IgA (sIgA) stimulation following the vaccine booster, the salivary IgG strongly tracked the serum IgG levels as recently reported^33^. It is possible that a sufficient concentration of salivary IgG could potentially block SARS-CoV-2 entry into oral epithelial cells^34^. Several studies also reported Ab avidity increased after booster vaccination against Wuhan and Omicron strains^31, 35, 36^. Ab avidity after the Wuhan-based booster vaccine also improved significantly overall in our study, though for the Wuhan and Delta strains were much higher than the Omicron. By comparison, the avidity index and the plasma Ab levels for Omicron following the booster were comparable to the Wuhan strain at six months following the two-dose vaccination before the booster. Our study is also unique in illustrating the improvement in the neutralizing Ab breadth after the third boost (T3) against Bat SARS-like coronavirus RsSHC014, representing a divergent member of the Sarbecovirus family. Interestingly, the Ab avidity is slightly stronger towards SHC014 than Omicron, suggesting that choice mutations in the RBD region drive escape from neutralizing antibodies.

Overall, our findings reinforce that repeated vaccine doses help the medium and low-responding vaccinees gain depth and breadth in plasma Abs to increase protection against highly evolving SARS-CoV-2 strains. When a new strain appears with substantial antigenic drift from the ancestral SARS-CoV-2 strain, the optimal strategy may rely on vaccines tailored to the new strains circulating among various populations. The currently distributed bivalent booster vaccines contain mRNA sequences for the spike protein of both ancestral and Omicron subvariants (BA.4/BA.5). However, uptake of the bivalent booster in the U.S population who were previously vaccinated is under 16% as of February 11th, 2023. Even among 65 and older U.S. residents, the bivalent booster uptake is under 41%, suggesting the need for new strategies to build confidence and promote the use of the SARS-CoV-2 vaccine^37^.

A recent CDC report recommends using evidence-based strategies to help increase vaccine confidence and uptake^38^. As a proof-of-concept, we utilized two US FDA-authorized blood- or saliva-based low lateral flow POCTs to detect inadequate SARS-CoV-2 humoral immunity in a cohort of healthy adults who showed variable neutralizing antibodies following 2-dose mRNA vaccination^3^. The negative POCT results were rapid (in under 20 minutes) and led to a definitive agreement on the absence or lack of functional Abs for the Wuhan, Delta, and BA.1 Omicron strains. Thus, POCTs can be great predictors of lacking nAb protection in people. However, developing POC Ab tests designed to match the circulating variants may be necessary to support an evidence-informed decision on receiving a booster or modified vaccine to protect against evolving and increasingly immune-evasive newer SARS-CoV-2 strains. As of Dec 2022, there are 46 vaccines approved for use in phase IV trials in various countries, and many counties have implemented more than one vaccine platform to use in people. These vaccines utilize different platforms, including protein subunit, virus-like particle, DNA, RNA, non-replicating viral vector, replicating viral vector, non-replicating viral vector, and inactivated and live-attenuated virus, and their efficacy differs against SARS-CoV-2 variants^39^. Depending on the circumstances, a negative SARS-CoV-2 POC Ab test result can be utilized to rapidly and economically inform those who can benefit from booster vaccination at the individual level, but also as a tool to assess herd immunity of the population at large as new variants emerge. POCTs can be regularly updated and rapidly deployed much more quickly than variant-matched SARS-CoV-2 vaccines.

Currently, several cost-effective SARS-CoV-2 POCTs are available to rapidly detect the viral protein (Antigen tests) or genetic material (molecular tests). At-home POC Ag tests, in particular, played a significant role in controlling the spread of the pandemic^40^. The tests facilitated early detection of virus infection and allowed individuals to self-isolate and seek early medical treatment. Additionally, POCTs offer the convenience of testing at home, do not require access to testing centers, and reduce demand on healthcare facilities by self-testing rather than at a clinic or hospital. Several authorized POCTs, such as those described in this manuscript, are also available to detect Abs using blood samples collected with a less painful fingerstick or saliva collected from non-invasive methods. However, usage of the POC Ab test is limited^41, 42^. A positive POC Ab test does not necessarily confirm that an individual is immune to COVID-19 or can’t transmit the virus to others, and our study does not oppose these recommendations. However, our findings elaborate on the potential utilization of POC Ab tests, ideally variant-matched, to inform a lack of neutralizing Abs, thereby supporting evidence-informed decision-making regarding the current and future booster vaccination.

A limitation of our longitudinal cohort study is that our participants only include primarily young, healthy adults and do not contain children, chronically ill, or immunocompromised subjects. Our feasibility study has yet to incorporate subjects after the bivalent booster vaccine, and the commercial POCTs utilized were based on the spike antigen of the Wuhan strain. Nevertheless, our study evaluated the depth and breadth of cellular and Ab responses in systemic and salivary compartments. We showed that vaccine boosting with the third dose enhances Ab titers, neutralization activity and ACE2 blockage, and heightens Ab avidity across heterogeneous subjects, whose response varied after the second dose. We also presented a proof-of-concept showing how LFA POCTs with appropriately updated RBD antigens can support evidence-based strategies to help increase vaccine confidence and uptake.

## Materials and Methods

### Clinical study and specimen collection

We enrolled a total of 237 healthcare providers working at the emergency department (ED) of George Washington University Hospital (GWUH). Emails for recruitment were sent to all GWUH ED HCP personnel through an ED staff listserv. Additionally, to reach those not on the listserv, notifications were sent out through GWUH’s ED nurse/technician scheduling system, and fliers were placed in break rooms with quick response (QR) codes connecting to patient sign-up forms. Participation days and times overlapped nurse and technician shift changes to encourage ongoing and off-going staff to participate. The clinical roles of study participants included physicians, advanced practice providers, nurses, and emergency department technicians. The study was approved by the George Washington University IRB#: NCR202406. All ED HCP personnel participating in this study provided written informed consent. All personnel who consented to participate were included in the study. Samples were collected in May/June 2020 (baseline, T0), January 2021 (after the first dose, T1), March 2021 (after the second dose T2a), July/August 2021 (after the second dose/before booster, T2b) and November/December 2021 (after the booster, T3). Not all participants provided specimens at every sampling point. Also, laboratory analyses were not performed if the vaccination dates were unknown or the booster was received before T2b sampling point. Venous blood samples were collected into an SST tube, refrigerated overnight to allow for serum separation, and stored at −80° C until laboratory analysis. Saliva samples were collected into a specialized device (Oracol S14 saliva collection device, Malvern Medical Developments, UK) according to the manufacturer’s instructions and stored at −80° C until laboratory analysis.

### Live-virus Neutralization assay

Neutralization assays were conducted using full-length live reporter virus constructs of Wuhan SARS-CoV-2 D614G (Sequence Aq. No MT020880), Delta SARS-CoV-2 B.1.617.2 (Aq. No OV116969.1), Omicron SARS-CoV-2 B.1.1.529 (EPI_ISL_6647961), and SHC014-Bat-CoV (Aq. no. KC881005.1) with the nano-luciferase (nLuc) reporter gene replacing ORF 7a in SARS-CoV-2 variants, and ORF 8 in SHC014 as previously published reports^17, 43, 44^. Assays were conducted in a modified manner from previously reported^17^. Heat-inactivated serum samples were initially diluted 1:20 and then serially diluted 5-fold down a 96-well plate (Corning 3799) in virus growth medium (1X MEM (Gibco 11095080), 5% FBS (Hyclone SH30070.03HI) and 1% Penn-Strep (Gibco 10378016)) before transfer to the BSL3 laboratory for assay completion. In the BSL3 laboratory, nLuc viruses were individually diluted in the virus growth medium, added in equal volume to serum dilution plates, and incubated for 1hr at 37℃, 5% CO_2_. The virus and serum dilutions were then added to duplicate columns of a 96-well black plate (Corning 3916) seeded one day prior with 2×10^4^ Vero C1008 cells per well for a final virus dilution of 800 Plaque Forming Units (PFU) per well, and incubated at 37℃, 5% CO_2_. After 20-24hrs, the virus was quantified with the Promega Nano-Glo Luciferase Assay system (N1130) with a Promega GloMax Explorer (GM3500). Inhibitory Dose 50 (ID_50_) titer was defined as the serum dilution at which the observed Relative Light Units (RLU) were reduced by 50% compared to Virus+ Cell and Virus-only control wells as determined by an Excel macro and analyzed using GraphPad Prism 9.3.1.

### HCoV Focus Reduction Neutralization Test

NL63, OC43 and 229E viruses were grown and tittered on LLC MK2 cells. To measure antibody mediate neutralization four-fold serial dilutions of human serum were mixed with ∼70 focus-forming units (FFU) of virus, incubated at 37°C for 1 h, and added to LLC MK2 monolayers in 96-well plates for 1 h at 37°C to allow virus adsorption. Cells were overlaid with 2% methylcellulose mixed with DMEM containing 5% FBS and incubated for 24 hours at 37⁰C. Media was removed and the monolayers were fixed with 5% paraformaldehyde in PBS for 15 min at room temperature, rinsed, and permeabilized in Perm Wash (PBS, 0.05% Triton-X). Infected cell foci were stained by incubating cells with polyclonal anti-NL63 (SinoBiological 40641-T62), anti-OC43 (SinoBiological 40643-T62), and anti-229E (MAB10938) for 1 h at 37°C and then washed three times with Perm Wash. Foci were detected after the cells were incubated with a 1:5000 dilution of horseradish peroxidase-conjugated goat anti-rabbit IgG for NL63 and OC43 and 1:5000 dilution of horseradish peroxidase-conjugated goat anti-mouse IgG (Sigma) for 1 h. After three washes with Perm Wash, staining was visualized by addition of TrueBlue detection reagent (KPL). Infected foci were then enumerated by CTL Elispot. FRNT curves were generated by log-transformation of the x axis followed by non-linear curve fit regression analysis using Graphpad Prism 8.

### Multiplex SARS-CoV-2 IgG and sIgA assays for oral fluid

Oral fluid IgG and sIgA to SARS-CoV-2 was measured with a multiplex immunoassay (MIA) as described previously^45^. Briefly, after thawing and centrifuging oral fluid for 5 min at 10,000 g, 10 μL supernatant were added to each microtiter well that contained 1000 coupled beads per bead set in 40 μL assay buffer (PBST with 0.1% BSA and sodium azide). Each multiplex IgG plate contained a blank well with assay buffer instead of sample for background correction, high and low SARS-CoV-2 IgG positive controls, and a SARS-CoV-2 IgG negative control in addition to an in-house SARS-CoV-2 IgG standard curve. Phycoerythrin-labeled anti-human IgG diluted 1:100 in assay buffer was used to detect the IgG signal in saliva (Jackson ImmunoResearch, 109-115-098). A monoclonal anti-human secretory component antibody, followed by a PE-labeled anti-mouse antibody was used to detect anti-SARS-CoV-2 sIgA. The plate was read on a Luminex MAGPIX instrument.

### T-cell receptor variable beta chain sequencing

Immunosequencing of the CDR3 regions of human TCRβ chains was performed using the immunoSEQ® Assay (Adaptive Biotechnologies, Seattle, WA). Extracted genomic DNA was amplified in a bias-controlled multiplex PCR, followed by high-throughput sequencing. Sequences were collapsed and filtered to identify and quantitate the absolute abundance of each unique TCRβ CDR3 region for further analysis as previously described^46–48^. The fraction of T cells was calculated by normalizing TCRβ template counts to the total amount of DNA usable for TCR sequencing. The amount of functional DNA was determined by PCR amplification and sequencing of several reference genes that are expected to be present in all nucleated cells.

### Mapping of SARS-CoV-2 TCRβ Sequences

TCR sequences from repertoires were mapped against a set of TCR sequences known to react to SARS-CoV-2. These sequences were first identified by Multiplex Identification of T-cell Receptor Antigen Specificity (MIRA)^49^. TCRs that react were further screened for enrichment in COVID-19 positive repertoires collected as part of immuneCODE^18^ compared to COVID-19 negative repertoires to remove TCRs that may be highly public or cross-reactive to common antigens. The individual response could be quantified by the number and frequency of SARS-CoV-2 TCRs seen post-vaccination. TCRs were further analyzed at the level-specific ORF or position within ORF based on the MIRA antigens.

### Recombinant protein antigens

The expression and purification of SARS-CoV-2 full-length spike ectodomain (16-1208 amino acids, Accession: P0DTC2.1), HaloTagged RBD (331-528 aa) and nucleocapsid antigens were as previously described^3, 50, 51^. The HaloTagged SARS-CoV-2 RBD antigens for bat SARS-like coronavirus RsSHC014, Delta SARS-CoV-2 B.1.617.2, Omicron SARS-CoV-2 B.1.1.529 were designed and expressed in mammalian Expi293 cells as described for RBD antigens^3, 50, 51^. RBD antigens were site-specifically biotinylated using HaloTag PEG-biotin ligand (Promega G8281), following the manufacturer’s protocol. The purified full-length ectodomain of the human coronavirus spike proteins (HCoV-NL63, 40604-V08B; HCoV-OC43, 40607-V08B, HCoV-229E, 40605-V08B, and HCoV-HKU1, 40606-V08B) were purchased from Sino Biological.

### Spike ELISA

Full-length spike ELISA was performed as described before^51^. Briefly, full-length spike protein at 2 μg/mL in TBS (pH 7.4) was coated in a high-binding microtiter plate (Greiner Bio-One 655061) and for 1 hour at 37°C, then blocked with blocking solution (3% milk in 0.05% TBST) for 1 hour at 37°C.Serum samples were serially diluted (1:33 – 1:8,100) in blocking solution and then added to the plate then incubated for 1 hour at 37°C. The plate was washed using BioTek 405 LS microplate washer, and horseradish peroxidase-conjugated secondary Goat Anti-Human secondary Ab IgG at 1:40,000 in 3% milk TBST blocking solution was added for 1 hour at 37°C. After washing the plate, 50 μL of 3,3′,5,5′-Tetramethylbenzidine (TMB) Liquid Substrate (Sigma-Aldrich T0440) was added, and absorbance was measured at 450 nm after stopping the reaction with 50 μL of 1 N HCl. The area under the curve (AUC) was calculated using GraphPad Prism.

### RBD ELISA

RBD ELISA was performed as described before^3^. High-binding microtiter wells were coated with Streptavidin (Invitrogen 434302) at 4 μg/mL in Tris-Buffered Saline (TBS) pH 7.4, incubated for 1 hour at 37°C, then blocked with Non-Animal Protein-BLOCKER™ (G-Biosciences 786190T) in TBS and incubated for 1 hour at 37°C. Serially diluted serum samples (1:33–1:8,100) were prepared in a 3% Bovine Serum Albumin solution (BSA) with TBST(TBS and 0.05% Tween-20) containing 3% BSA and 1 μg/mL biotinylated RBD antigen, then incubated in a non-binding 96-well plate for 1 hour at 37°C. Following incubation, the diluted serum was added to the microtiter assay plate where it was incubated for 15 minutes at 37°C. After washing, horseradish peroxidase-conjugated secondary Goat Anti-Human secondary IgG Ab at 1:40,000 dilution in 3% milk TBST was added, and the plate was incubated for 40 minutes at 37°C. After washing, 3,3′,5,5′-Tetramethylbenzidine (TMB) Liquid Substrate (Sigma-Aldrich) was used to develop the color. Absorbance was measured at 450 nm after stopping the reaction with 50 μl of 1 N HCl. A 3-fold serially diluted anti-SARS-CoV-2 Spike RBD monoclonal Ab (Abeomics, ABMX-002) was used in every assay plate. A standard curve obtained from the spike RBD mAb was used to define the concentration of Abs in the clinical samples (1 binding Ab unity (BAU) = 1 ng/ml of the mAb).

### Nucelocapsid ELISA

The SARS-CoV-2 nucleocapsid IgG ELISA was performed as previously described^3^. High-binding microtiter wells (Greiner Bio-One 655061) were coated with 3 μg/mL anti-MBP mAb (E8032, New England Biolabs) and incubated for 1 hour at 37°C, then blocked with blocking solution (3% non-fat powdered milk in Tris-Buffered saline and 0.05% Tween-20). After washing the plate with TBS containing 0.2% Tween-20, 2 μg/mL MBP-fused full-length nucleocapsid or MBP protein in blocking solution was added to respective wells and incubated for 1 hour at 37°C. After washing the plate, heat-inactivated serum at a 1:40 dilution was added and incubated for 1 hour at 37°C. The plate was washed, and alkaline phosphatase-conjugated secondary goat anti-Human anti-IgG (Sigma A9544) was added to the wells at 1:2,500 dilution. Absorbance was measured at 405 nm after adding the SIGMAFAST p-Nitrophenyl phosphate substrate (SigmaN2770). Appropriate control sera were included in the study. The nucleocapsid binding signal for each serum was calculated by subtracting the absorbance of the background signal obtained from MBP wells.

### Avidity index of IgG Abs

High-binding microtiter wells were coated with streptavidin at 4 μg/mL in tris-buffered saline (TBS, pH 7.4) for 1 hour at 37°C, then blocked with Non-Animal Protein-BLOCKER™ (G-Biosciences) in TBS and incubated for 1 hour at 37°C. The plate was washed 3 times with wash buffer (TBS containing 0.2% Tween 20), and a TBST buffer (TBS and 0.05% Tween-20) containing 3% BSA and biotinylated RBD at 1 μg/mL was added and incubated for 30 minutes at 37°C. After the plate was washed, heat-inactivated serum samples at 1:900 in TBST containing 3% BSA were added and incubated for 1 hour at 37°C. After washing the plate with wash buffer, wells were incubated with and without 4 M urea containing TBST + 3% BSA for 25 minutes at 37°C. Urea solution was dumped and washed three times, and horseradish peroxidase-conjugated secondary Goat Anti-Human secondary IgG Ab at 1:40,000 dilution in 3% milk was added. The plate was incubated for 40 minutes at 37°C. 3,3′,5,5′ Tetramethylbenzidine (TMB) Liquid Substrate (Sigma-Aldrich) was used for signal development, and absorbance was measured at 450 nm after stopping the reaction with 50 μl of 1 N HCl. The avidity index was calculated as the absorbance in the presence of urea/absorbance in the absence of urea.

### Multiplex surrogate neutralization assay

A multiplexed Meso Scale Discovery (MSD) immunoassay (MSD, Rockville, MD) was used to measure the ACE2 blocking Abs to SARS-CoV-2 Wuhan and other variants, including Alpha (B.1.1.7), Beta (B.1.351), Delta (B.1.617.2), IHU(B.1.640.2) and Omicron subvariants (BA1, BA2, and BA3), using the MSD V-PLEX SARS-CoV-2 Panel 25 as previously described^3^. Briefly, plates were blocked with MSD Blocker A for 30 minutes and washed thrice. Then the reference standard, controls, and heat-inactivated samples diluted 1:100 in the diluent buffer were added. Plates were incubated at room temperature for 1 hour with shaking at 700 rpm. MSD SULFO-tag conjugated ACE2 (0.25μg/ml) was added and incubated for 1 hour at room temperature with shaking. Plates were washed and read with a MESO QuickPlex SQ 120 instrument. ACE2 blocking activity was calculated using the equation: ((1 – Average Sample ECL Signal / Average ECL signal of the blank well) x 100).

### LFA Assay with Blood samples

Cellex™ qSARS-CoV-2 IgG/IgM Rapid test was performed following the manufacturer’s instructions. Briefly, 10 µL of blood sample was dispensed into the center of the sample disposal, followed by two drops of sample diluent provided by the kit were added to the sample disposal. The cassette was placed horizontally on a clean, flat surface, and the reaction was left to develop at room temperature for 20 min. The color changes in the test and control lines at 20 minutes were read by the naked eye, and a photograph was taken for record. A sample was considered positive if both test IgG or IgG+IgM and control line (C) were visible, negative if only “C” line was visible and there was no test line. The test was invalid if the “C” line failed to develop.

### LFA Assay with Saliva samples

CovAb™ SARS-CoV-2 Ab Rapid Test was performed with saliva samples described by manufacturer instructions on a clean, flat surface. Briefly, five drops of saliva sample diluted with the buffer provided by the manufacturer’s kit at a 1:1 were transferred to the sample well, allowing the reaction to run for 15 min. The visible test and control lines were read at the end of 15 minutes, and a photograph was taken for record. A sample was considered positive if both test and control lines were visible and negative if only a control line was visible and the test line was absent. The test was invalid if the “C” line failed to develop.

## Data Availability

All data produced in the present study are available upon reasonable request to the authors

## Acknowledgements

This work was supported by the NCI Serological Sciences Network (4U54CA260543 to RSB and LP; U01CA260469 to CDH; U01CA260541 and U01CA260541-02S1 to AMD and JDB). The funders had no role in study design, data collection, decision to publish, or preparation of the manuscript. We thank the health care providers and staff at the George Washington University Hospital Department of Emergency Medicine for their contributions to this work. The authors are grateful for the in-kind support of Adaptive Biotechnologies, Inc. for the immunoSEQ assay. We acknowledge Cellex, NC, USA, for donating Cellex™ qSARS-CoV-2 IgG/IgM Rapid test kits for this study. TM is grateful for the support of the St. Laurent Institute and The Ulvi and Reykhan Kasimov Family.

## Supplementary Information

**Supplementary Figure 1.**
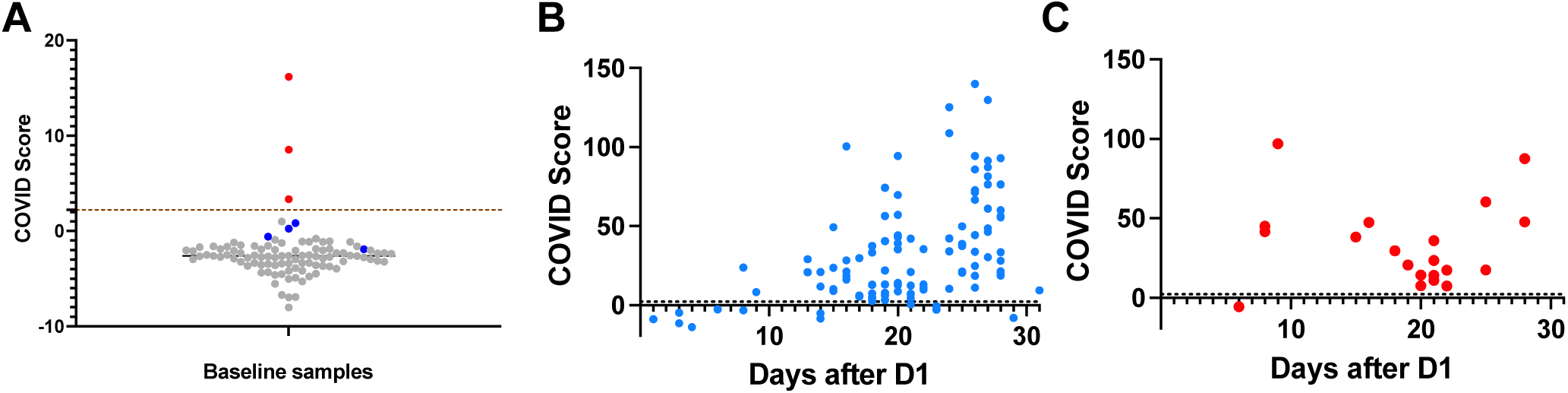
TCR COVID Score reliably detects following SARS-CoV-2 infection and vaccination. **(A)** Analysis of SARS-CoV-2 TCR-β sequences prior to vaccination. Blood samples from 99 subjects that included 7 PCR positives were analyzed. Samples from 3 of 7 COVID-positives were collected >8 days after PCR confirmation (red), and the remaining samples from 4 COVID-positives were collected <4 days after PCR confirmation (blue). A COVID score threshold of >2.23 was applied to classify the subjects as positive or negative. Percent positivity at each time-point is shown. COVID Score is 100% specific and sensitive with pre-vaccine samples collected >8 days after PCR confirmation. Longitudinal analysis of SARS-CoV-2 TCR-**β** sequences following dose 1 of the mRNA vaccine in (B) Naive and (C) previously-infected subjects. Blood samples collected between 1 and 31 days from 111 naïve individuals who received an primary mRNA vaccination were analyzed. Of the seven samples collected <8 days, one was positive by the COVID Score cut-off (2.23). Of the remaining 104 samples collected nine days after the first vaccine dose, 97/104 were positive by COVID Score, yielding a sensitivity of 93.7% among naive subjects after dose 1. Blood samples were analyzed from 20 previously infected subjects who received a primary mRNA vaccination. Excluding a sample collected on day six post-dose 1, the remaining 19 samples collected between 8 and 28 days were positive, yielding a sensitivity of 95% among previously infected subjects after dose 1.

**Supplementary Figure 2.**
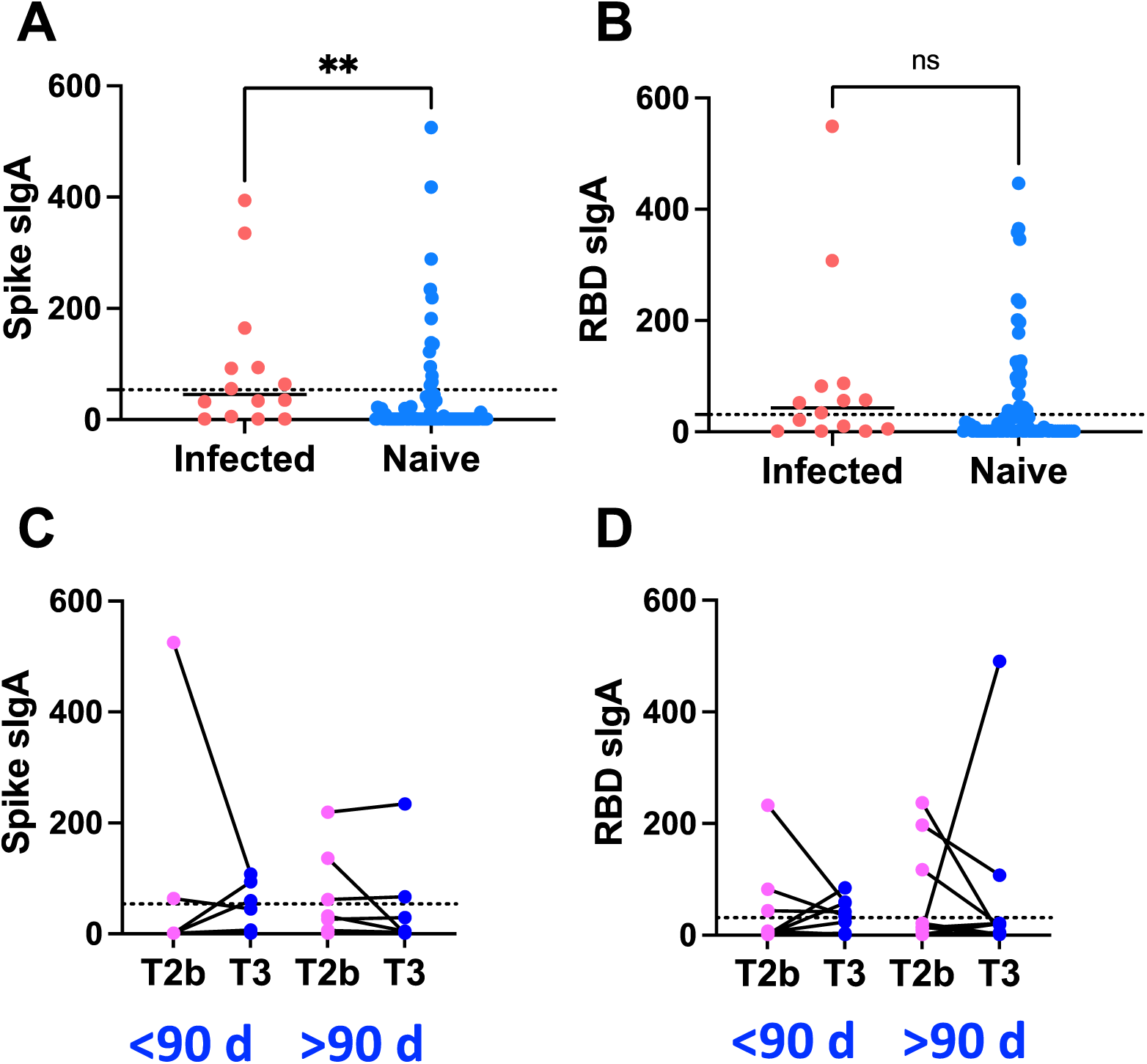
Absence of salivary secretory IgA (sIgA) Abs following the mRNA vaccine booster. Comparison of sIgA levels in infected-vaccinated and naive-vaccinated subjects against (A) Spike and (B) RBD antigens. A multiplex Luminex assay was used to assess sIgA levels in saliva samples (n=14, infected vaccinated and n=77, naive-vaccinated) collected at pre- and post-booster. A Mann-Whitney non-parametric test to determine if sIgA levels differ between the groups showed that the infected vaccinated group had a modestly higher anti-spike sIgA level than naïve boosted group (p=0.0021). However, the difference between these groups was insignificant in anti-RBD sIgA levels. Assessment of sIgA levels in paired pre- and post-booster samples against (C) Spike and (D) RBD antigens. The sIgA levels to spike and RBD in post-booster samples collected <90 days from 15 subjects or collected >90 days post-booster from 9 subjects were compared to their respective pre-booster levels. As indicated by the Wilcoxon-non-parametric paired test, the sIgA levels in pre- and post-booster paired samples are comparable, suggesting a lack of evidence for sIgA development after the third booster shot. The plot’s dotted line represents the sIgA assay thresholds for Spike sIgA (53.6 MFI, BSA background subtracted) and RBD (31 MFI, BSA background subtracted), determined based on the reference panel, was shown.

**Supplementary Figure 3:**
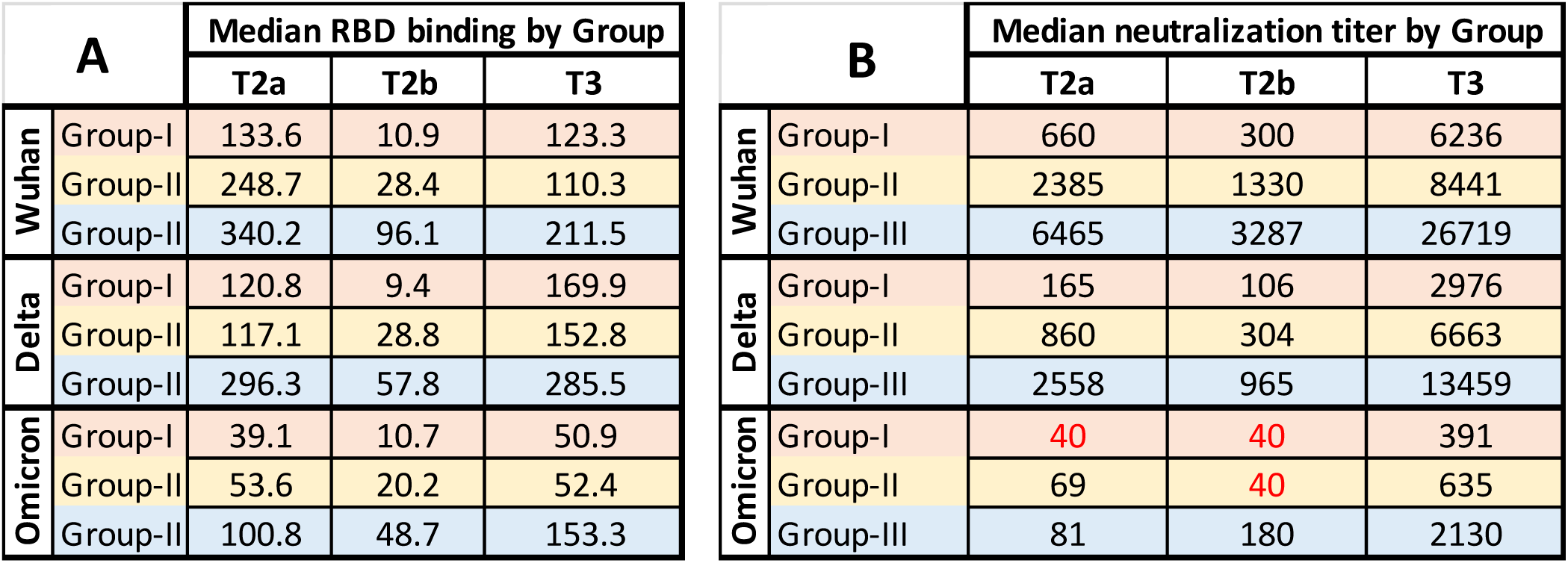
Dynamics of binding and neutralizing antibody response before and after the booster. Median RBD binding Abs (A) and neutralizing Abs (B) to the Wuhan, delta, and omicron variants in each of the three groups (I, II, and III) at 4-9 weeks after dose-2 (T2a), 19-36 weeks after dose-2 (T2b) and 3-18 weeks after dose-3 (T3). Group I (n=11) had a weak neutralizing Ab response after complete vaccination. Group II (n=8) had a robust neutralizing Ab response after dose-1 that declined after dose-2. Group III neutralizing Abs rose after dose-1 and after dose-2. Median RBD binding and neutralizing Abs decreased between T2a and T2b and sharply raised after the third booster dose in all three groups. The Group I binding and neutralizing Abs levels were the lowest at T2b and sharply improved after the third booster than Groups II and III. Notably, before receiving the third booster dose, omicron-neutralizing Ab titers were below the limit of the assay for Groups I and II (40, red).

**Supplementary Figure 4.**
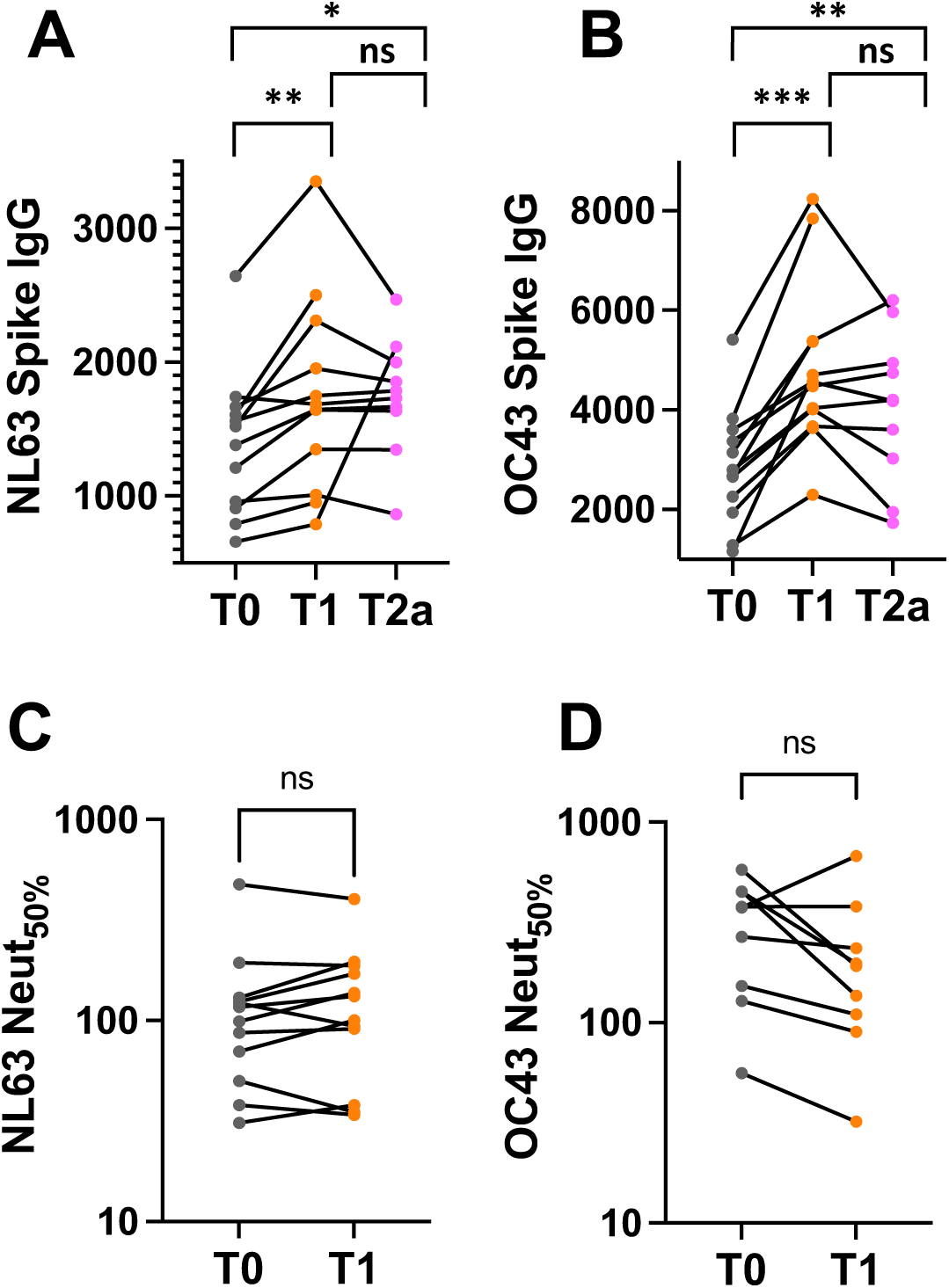
HCoV CR reactive Abs induced following mRNA vaccine lacks cross-neutralizing activity. Analysis of spike IgG Abs to (A) NL63 and (B) OC43 prior to vaccination and after doses 1 and 2. Binding IgG Abs were measured by ELISA using serially diluted sera from 12 subjects collected at pre-vaccine, after dose 1 (2-4 weeks), and after dose 2 (5-9 weeks) and expressed as AUC. Analysis of neutralizing Abs to (C) NL63 and (D) OC43 at pre-vaccine and after doses 1 and 2. Wilcoxon-non-parametric paired test was performed to determine if Ab response to NL63 and OC43 between baseline and after doses differ. The test result showed that even though the spike binding Ab to seasonal CoVs is elevated between baseline and after vaccination, they do not effectively contribute to cross-neutralization. The p-value summary from the Wilcoxon test is shown above the scatterplot. p < ∗∗∗0.0005; ∗∗0.0020; ∗0.0332; ns, >0.075.

**Supplementary Figure 5.**
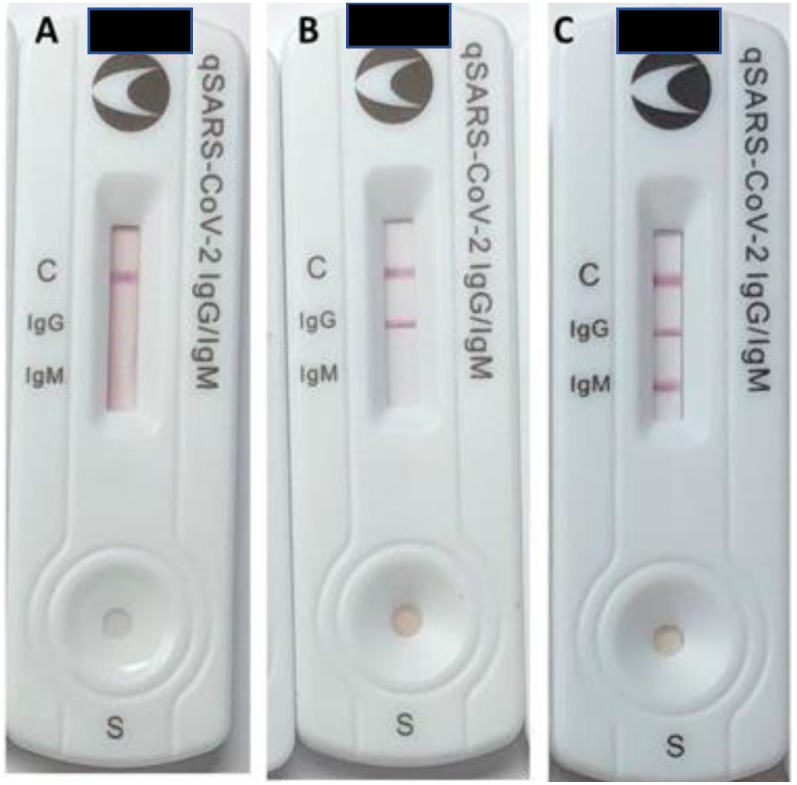
CELLEX LFA test results with serum samples for (A) valid negative (B) valid IgG positive and (C) valid IgG+IgM positive readings.

**Supplementary Figure 6.**
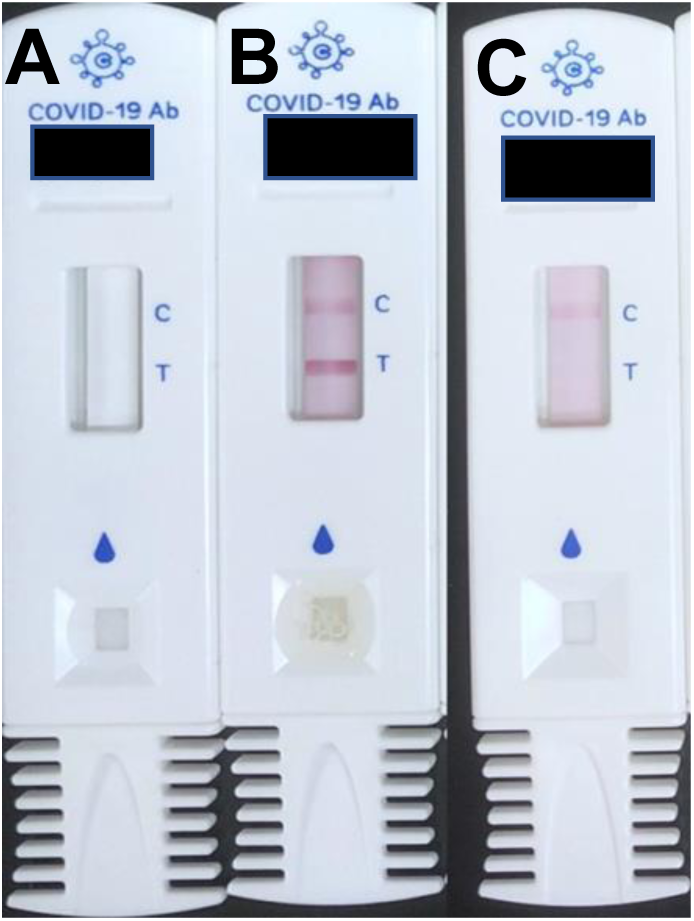
COVAB LFA test results with salivary samples for (A) invalid (B) valid positive and (C) valid negative readings.

